# Virtual Reality for Pediatric Trauma Education - A Face and Content Validation Study

**DOI:** 10.1101/2024.02.16.24302807

**Authors:** Said Ashkar, Fabio Botelho, Shreenik Kundu, TJ Matthews, Elena Guadgano, Jason Harley, Dan Poenaru

**Author notes:** **Corresponding author** Fabio Botelho, MD, MSc, Harvey E. Beardmore Division of Pediatric Surgery, Montreal Children’s Hospital, 1001 Decarie Boulevard, Montreal, QCH4A3J1, Canada. Said Ashkar and Fabio Botelho are both first-authors.

## Abstract

**Purpose:** Pediatric trauma is a leading cause of death and disability among children. While trauma education can improve these outcomes, it remains expensive and available only to a few providers worldwide. Innovative educational technologies like virtual reality (VR) can be key to democratizing trauma education. This study, therefore, evaluates the face and content validity of a VR platform designed to enhance pediatric trauma skills. Specifically, we seek to determine whether the platform effectively presents an injured child and comprehensively covers the essential tasks to successfully treat them within a trauma team.

**Methods:** Physicians were invited to test a VR platform simulating a child with blunt head and truncal trauma. After the simulation, they filled out surveys assessing the face and content validity of the scenario, including their opinions on the realism, interaction, ease of use, and the educational content of the platform. Additionally, they completed a cybersickness questionnaire. Demographic data were also collected, including age, gender, country of medical education, and previous experience with VR. A descriptive analysis was performed.

**Results:** Eleven physicians graduated from eight different countries tested the VR platform. Most (87%) found it valuable, and 81% preferred using it over high-fidelity mannequins for training purposes. The platform received more favorable evaluations for non-technical skills training (median: 5, IQR: 5.0 to 5.0) than for technical skills (median: 4, IQR: 3.0 to 5.0). Regarding cybersickness, 73% of the participants reported experiencing any or minimal discomfort during the simulation, and none needed to stop the test due to discomfort.

**Conclusion:** Our initial validation of a VR platform designed for pediatric trauma education was positive. Participants endorsed VR and its potential to enhance performance, particularly in non-technical skills. Encouraged by these results, we will proceed with feasibility and implementation studies, comparing VR to high-fidelity mannequins.

## I. Introduction

Trauma continues to be a critical health problem worldwide, particularly in pediatric populations, claiming the lives of a million children annually.[1] According to the World Health Organization, trauma has been a leading cause of child mortality in 204 countries for the past three decades.[2] Despite significant advancements in trauma care during this period, mortality rates have remained high.[1]

Errors in care persist and are often linked to poor communication and decision-making among trauma team members.[3,4] Providing training in non-technical skills, such as situational awareness, leadership, and teamwork, can change this reality.[5,6] Non-technical skills foster efficient collaboration during challenging and stressful environments, enhancing the quality of care.[7,8] However, the methods for teaching and assessing these skills are not standardized, revealing notable discrepancies in pediatric trauma training approaches across different settings.[8,9]

In low- and middle-income countries (LMICs), pediatric trauma care is also hindered by resource limitations, a shortage of expert providers, and the scarcity of training programs.[10,11] The use of advanced technologies for trauma education, like high-fidelity mannequins, has unfortunately exacerbated inequities. The current courses are prohibitively expensive and, consequently, not viable for widespread implementation,[12,13] remaining accessible only to a limited part of the population.[10,14,15]

Virtual reality (VR) has emerged as an innovation in healthcare that can help address these gaps. [16] By creating engaging and realistic clinical environments, VR can help healthcare professionals refine their skills in a setting that imitates real-world demands.[17,18] In LMICs, VR also presents a potential opportunity to democratize access to pediatric trauma education without incurring heavy expenses on physical equipment.[19] Moreover, with built-in, instant feedback systems, platforms with automatic feedback can also facilitate debriefing, enhancing the learning experience.[17]

To our knowledge, only one VR platform specific to the pediatric emergency room has been documented in the literature. [20] Abulfaraj et al. randomly assigned pediatric and emergency medicine interns to their single-user VR platform or to a high-fidelity mannequin for a seizure simulation. The time-to-critical actions, such as oral suction and oxygen administration, demonstrated no statistically significant differences between the VR and mannequin groups. Notably, limited prior experience with VR gaming was reported, and most participants displayed a significant increase in subjective confidence after the simulation sessions.

The literature has also documented single-user VR platforms tailored for adult trauma scenarios, which have the potential to reduce the cost of training. [17,21,22] Cohen et al. designed a 3-scenario adult trauma VR simulation for which they found a high level of acceptance and perceived realism among participants, with 96% considering the virtual environment valuable. [23]

In another study evaluating a single-user VR for ATLS, participants were satisfied with the platform and learning experience.[24] Furthermore, the results illustrated adequate construct validity, with attending physicians achieving the highest decision accuracy and the lowest mortality rate compared to trainees.

Cybersickness is the negative side of VR. It is characterized by symptoms akin to motion sickness, such as dizziness and nausea, experienced in virtual environments without actual physical movement.[25,26] The reported incidence of cybersickness varies widely in the literature, with some studies indicating minimal effects (less than 10%) while others suggest rates as high as 80%.[20,25,27] The incidence of cybersickness is influenced by numerous factors, including the users’ roles and actions in the virtual simulation and the duration of exposure.[25,27] An initial evaluation of cybersickness is instrumental in guiding the development of further scenarios, as the presence of symptoms does not necessarily require abandoning the task, being the reason we decided to include it as a measure. Such symptoms rather inform decisions to potentially restrict usage time or decrease the need to move (virtually) during the assessment.

This study aims to assess the face and content validity of a multi-user virtual simulation platform for pediatric trauma training called PeTIT VR - Pediatric Trauma Innovative Training in Virtual Reality. (*Figure 1*) The platform accommodates up to five users concurrently – constituting four trainees and an instructor – enabling them to enter into a 3D environment to practice the initial trauma assessment, comprising primary and secondary surveys. All virtual trauma bay assets and interactions were crafted with input from pediatric trauma experts to maximize realism and educational value. Scenarios are set to evolve dynamically in response to trainee actions, and quantitative and qualitative performance feedback is automatically provided at the end to facilitate debriefing. A screen appears, informing students about the tasks they completed or missed (for example, checking the airway and starting fluids) and also displays the time it took for the user to accomplish each task.

**Figure 1.**
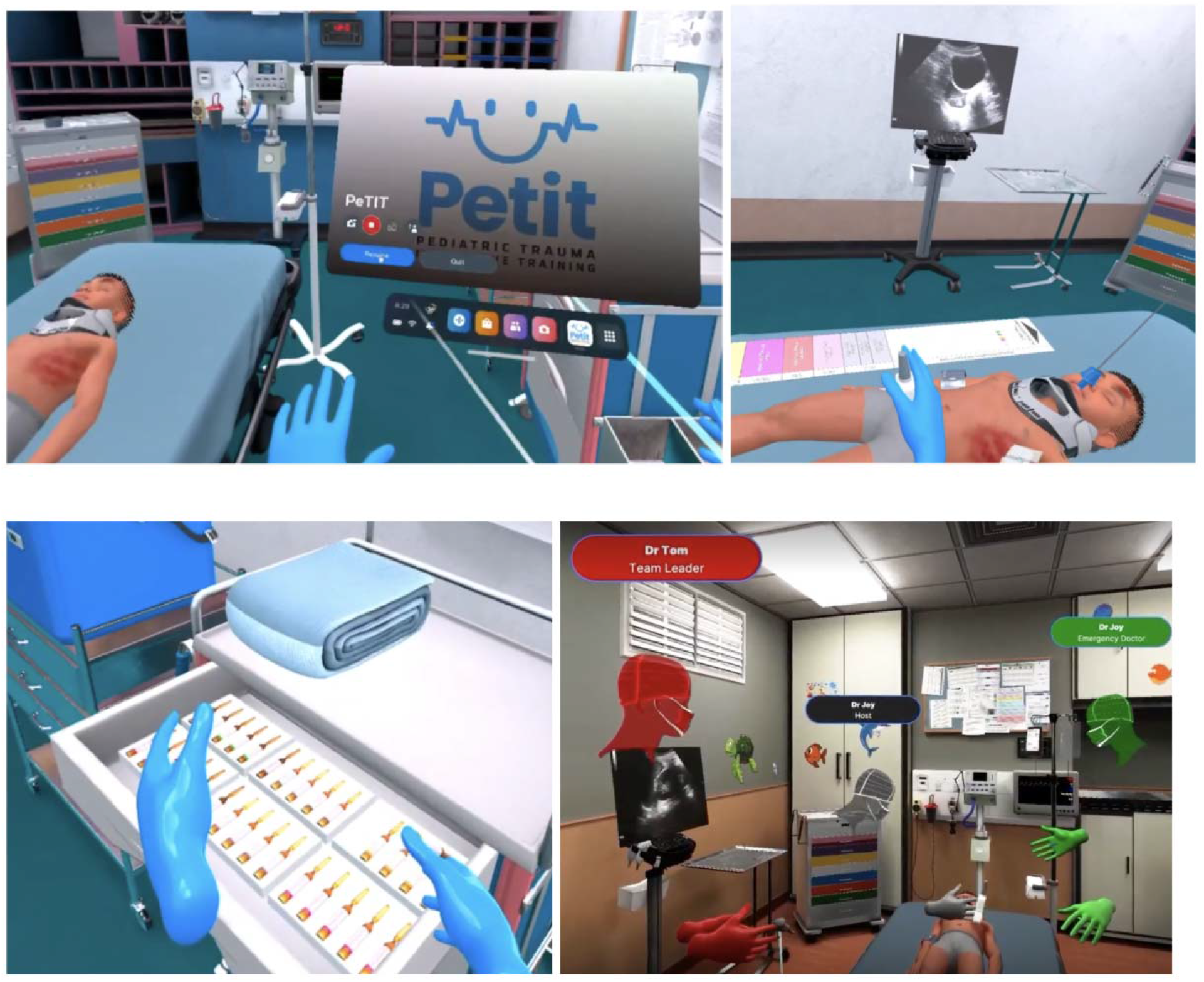
PeTIT VR

This study seeks to examine face and construct validity and represents the initial phase of an extensive research agenda. The platform has been used for research purposes and is not currently commercially available.

## II. Methods

### Participants

Eleven physicians participated in the study *(Table 1)*. Of these, five were male (45%), and six were female (55%), with a median age of 35 years (range 29 - 54 years). Participants had graduated from eight countries, including Canada, Brazil, Argentina, Afghanistan, Iran, the United States of America, Sudan, and Nigeria *(Figure 2)*. Among them, five were staff pediatric surgeons (45%), three identified as residents (27%), one as a pediatric emergency staff (9%), one as an emergency physician (9%), and one as a general practitioner (9%). Approximately one-third (36%) of them had prior experience with VR.

**Table 1.** Demographic distribution of the participants.

|  | No. participants (%) |
| --- | --- |
| ***Age (y)*** |  |
| 25-30 | 1 (9) |
| 31-35 | 6 (54) |
| 36-40 | 2 (18) |
| > 41 | 2 (18) |
| ***Specialty*** |  |
| Pediatric surgery | 8 (72) |
| Emergency medicine | 2 (18) |
| General practice | 1 (9) |
| ***Gender*** |  |
| Male | 5 (45) |
| Female | 6 (55) |
| ***Country of education*** |  |
| Afghanistan | 1 (9) |
| Argentina | 1 (9) |
| Brazil | 1 (9) |
| Canada | 4 (36) |
| Iran | 1 (9) |
| Nigeria | 1 (9) |
| Sudan | 1 (9) |
| USA | 1 (9) |

**Figure 2.**
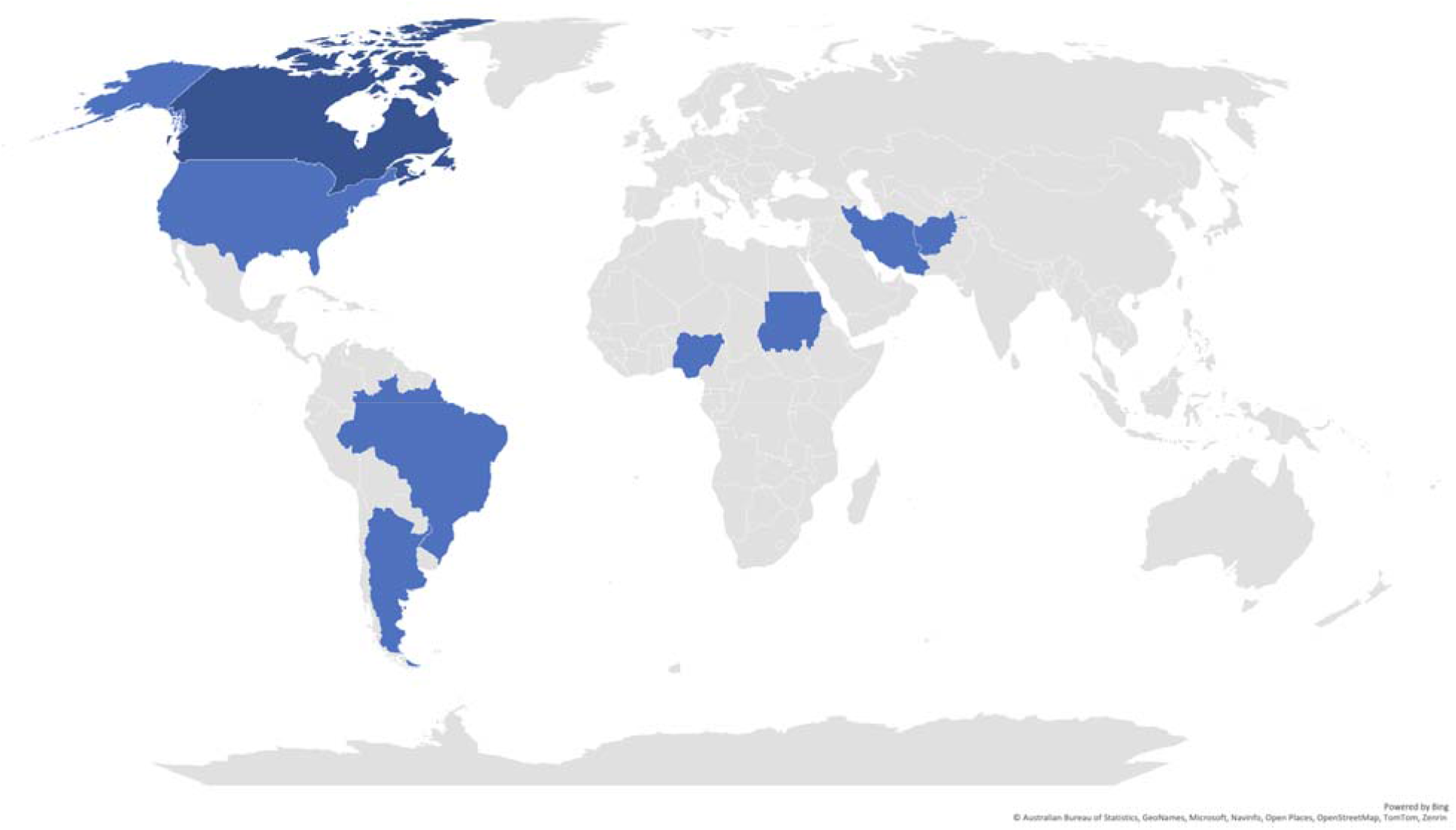
Geographic distribution of medical institutions where participants completed their medical training

### Measures

To assess the face and content validity of our platform, participants were invited to complete two questionnaires based on prior VR research. [20,21] Given the absence of validated questionnaires for trauma education, the authors collaboratively adapted the questions for the field. Two independent reviewers subsequently examined the survey for readability and potential for misinterpretation. Additionally, a third survey based on the Technology Acceptance Model was administered to collect preliminary feedback on user acceptance of the technology. This survey included questions deemed most pertinent to our platform and trauma education by the authors. We also assessed cybersickness using the methodology described by Kim et al.[22]

#### Face Validity

The face validity questionnaire comprised five questions evaluating the VR platform’s realism and appropriateness compared to real-life situations or mannequins. Questions included whether “*the anatomic structures of the patient appear realistic*,” “*the interactions (e*.*g*., *pulse examination, capillary refill, auscultation) are realistic*,” “*the virtual patient meets my expectations*,” and “*the experience with the virtual patient is better than with high-fidelity mannequins*.” Responses were recorded on a five-point Likert scale, ranging from “strongly disagree” to “strongly agree.”

#### Content Validity

A similar five-point Likert scale questionnaire was employed to assess content validity. It evaluated the accuracy of the primary and secondary surveys and their effectiveness in fostering both technical and non-technical skills. The relevance of the platform for training diverse learners, including medical students, residents, and staff physicians, was also reviewed.

#### Technology Acceptance

Four questions from the Technology Acceptance Model survey were selected and adapted to assess ease of use (“*I found PETIT VR easy to use*”), overall experience (“*my overall experience with PETIT VR was positive*”), likelihood of recommendation (“*I would recommend PETIT VR for pediatric trauma training*”), and potential to increase interest in trauma training (“*PETIT VR can increase my interest in pediatric trauma training*”). These were also answered using a five-point Likert scale to maintain survey uniformity and simplify the response process.

#### Cybersickness

The Virtual Reality Sickness Questionnaire (VRSQ) measures symptom intensity on a scale. [22] Symptoms assessed include general discomfort, fatigue, eyestrain, difficulty focusing, headache, fullness of head, blurred vision, dizziness (with eyes closed), and vertigo. We adapted the scale to a five-point Likert format for consistency with the other surveys, with 0 indicating no symptoms, scores of 2 and 3 categorized as some discomfort, and 4 and 5 as significant discomfort.

#### Qualitative Feedback

Participants were invited to provide additional comments in a free-text format for aspects not covered by the surveys.

### Procedures and Simulation Environment

The initial VR scenario portrayed an injured child post-road traffic collision, with head trauma, a tension pneumothorax, and abdominal bleeding. Critical decisions included intubation, chest tube insertion, fluid and blood transfusion initiation, and consulting the general surgery team.

The scenario lasted approximately 15 minutes. Participants engaged in the scenario twice, first as the trauma team leader, to enhance leadership skills, and then with full interaction with the virtual tools and team roles.

#### Part 1

Participants assumed the role of the trauma team leader, while research team members enacted other roles such as nurse, respiratory therapist, or emergency physician. The leader was positioned to oversee the room and lead and monitor team dynamics. They could also perform any tasks if team members were unable to do. These tasks could be talking to the patient and performing a physical exam that includes auscultation, checking for oral secretions, heart sounds, pulse via controller vibrations, and pupil reaction to light. Text prompts provided feedback for non-haptic physical exam components, like abdominal palpation. The patient reacted to pain and vocal stimuli and displayed physiological changes mirroring real-life scenarios.

#### Part 2

After the leadership role, participants explored all the platform’s tools, including the Broselow tape, intubation equipment, and the Focused Assessment with Sonography for Trauma (FAST) exam. This part lasted an additional 10 to 15 minutes.

After the testing session, participants completed the online surveys.

### Analysis

Descriptive analyses were conducted on the users’ answers. Results were summarized using medians and interquartile ranges. Comments from participants were also included in the Results section. For analyses, we utilized Microsoft Excel, version 16.73. The analyses were conducted in STATA version 18.

### Ethical considerations

The study was approved by the McGill University Health Centre, Research Ethics Board (#2024-10024). Participants authorized data collection through the online form.

## III. Results

### Face and content validity

The self-reported face validity and technology acceptance results indicate a positive reception of the VR platform among participants (*Table 2 and 3*). Most of them found the anatomic structures of the virtual patient realistic (median: 5, IQR: 4.0 to 5.0) and could identify injuries and conduct the physical exam using the platform (median: 5, IQR: 4.0, 5.0). Some interactions such as pulse assessment, capillary refill, and auscultation were rated slightly lower (median: 4, IQR: 4.0 to 5.0), but still positive. The virtual patient’s ability to meet trainees’ expectations was highly rated (median: 5.0, IQR: 5.0 to 5.0). VR was considered a better platform than high-fidelity mannequins (median: 4, IQR: 4.0 to 5.0). The *ease of use* was noted to be high (median: 4.0, IQR 4.0 to 4.0), and the participants unanimously acknowledged the platform’s potential to increase their interest in pediatric trauma training (median: 5; IQR: 4.0 to 5.0). The high willingness to recommend the platform (median: 5, IQR: 4.0 to 5.0) underscores its perceived utility.

**Table 2.** Face validity survey.

| Face validity statements | Median (IQR) |
| --- | --- |
| The anatomic structures of the patient appear realistic | 5 (4.0, 5.0) |
| The interaction (for example, pulses exam, capillary refill, auscultation) is realistic | 4 (4.0, 5.0) |
| I could identify injuries and abdominal exams using the platform | 5 (4.0, 5.0) |
| The virtual patient meets my expectations | 5 (4.0, 5.0) |
| The experience with the virtual patient is better than the high-fidelity mannequins (if you had experience with high-fidelity mannequins before) | 4 (4.0, 5.0) |

**Table 3.**
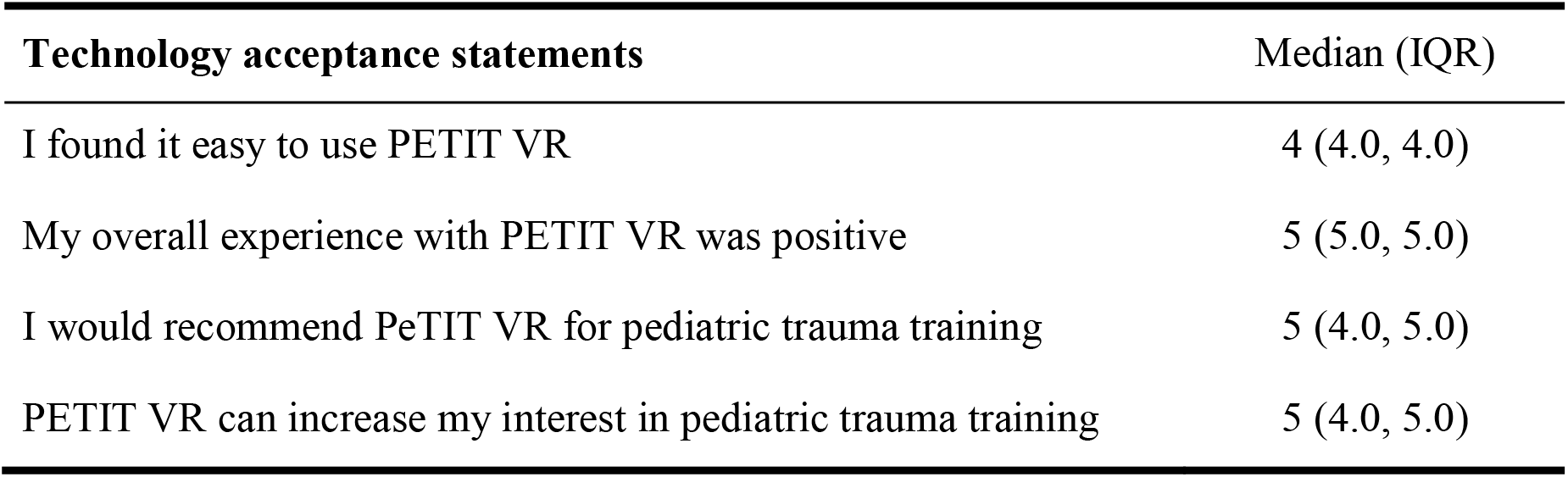
Technology acceptance survey.

| Technology acceptance statements | Median (IQR) |
| --- | --- |
| I found it easy to use PETIT VR | 4 (4.0, 4.0) |
| My overall experience with PETIT VR was positive | 5 (5.0, 5.0) |
| I would recommend PETIT VR for pediatric trauma training | 5 (4.0, 5.0) |
| PETIT VR can increase my interest in pediatric trauma training | 5 (4.0, 5.0) |

The potential of the VR platform in fulfilling pediatric trauma training requirements is detailed in *Table 4*. The median scores for all seven criteria ranged from 4.0 to 5.0, indicating an overall agreement (“agree” and “strongly agree” categories). Participants found the VR platform more advantageous for training non-technical skills (median: 5, IQR: 5.0 to 5.0) than technical skills (median: 4.0, IQR: 3.0 to 5.0). Testers reported that the platform could benefit training across various professional levels, including medical students, residents, and staff. Additionally, the ratings for the usefulness of the VR platform in primary and secondary surveys were also similarly high.

**Table 4.** Participant responses to the content validity questionnaire.

| Content validity statements | Median (IQR) |
| --- | --- |
| The current model is useful for training pediatric trauma primary survey | 5 (4.0, 5.0) |
| The current model is useful for training pediatric trauma secondary survey | 5 (4.0, 5.0) |
| The current model is useful for training technical skills (intubation, chest tube, FAST exam) | 4 (3.0, 5.0) |
| The current model is useful for training non-technical skills (leadership, communication, teamwork, decision-making) | 5 (5.0, 5.0) |
| The current model is useful for training medical students | 5 (5.0, 5.0) |
| The current model is useful for training residents | 5 (4.0, 5.0) |
| The current model is useful for training staff | 5 (4.0, 5.0) |

### Cybersickness

The cybersickness results revealed that 36% of participants experienced no issues with the VR platform, 36% reported minimal symptoms, and 27% experienced significant discomfort. A closer examination of these issues (*Table 5*) showed that the primary complaints were *difficulty focusing, blurry vision*, and *eye strain*.

**Table 5.** Cybersickness associated with the VR platform.

| Cybersickness parameters | No. participants (%) |  |  |
| --- | --- | --- | --- |
|  | None | Some | Significant |
| General discomfort | 7 (67) | 3 (27) | 1 (9) |
| Fatigue | 9 (82) | 2 (18) | 0 (0) |
| Eye strain | 7 (67) | 4 (36) | 0 (0) |
| Difficulty focusing | 5 (45) | 6 (55) | 0 (0) |
| Headache | 10 (91) | 1 (9) | 0 (0) |
| Fullness of head | 10 (91) | 1 (9) | 0 (0) |
| Blurry vision | 8 (73) | 1 (9) | 2 (18) |
| Dizzy (with eyes closed) | 10 (91) | 1 (9) | 0 (0) |
| Vertigo | 10 (91) | 1 (9) | 0 (0) |

### Qualitative feedback

The qualitative feedback highlighted the VR platform’s perceived realism and user-friendliness. All the comments received are registered below.

Users expressed satisfaction with the platform’s design, with one user noting, “*Excellent tool-very realistic, helpful*,” and another confirming the platform’s *ease of use*, stating, “*The system was easy to use and intuitive, and the case was comprehensively reproduced in the VR system*.” This positive sentiment extended to the platform’s potential in training for non-technical skills, as one physician highlighted: “*Great VR simulation, Pros: a good way to get training for CRM [Crew Resources Management]/non-medical objectives, no need to displace yourself to a sim center for training*.”

Users also pinpointed specific limitations of VR, particularly in practicing technical skills that rely heavily on haptic feedback. One participant remarked, “*I do not think it is an ideal platform to practice technical skills given the limitations*… *to become comfortable with procedures like intubation or chest tube, you have to ‘feel it’ to develop muscle memory*.*”* Echoing this sentiment, another noted the disadvantages of procedural training: “*Cons: Not a good way to test procedural skills as a lot is physical memory, therefore, (you) need to touch chest tubes, etc*.”

Visual clarity was another concern raised, with a user indicating a potential area for improvement: “*I was not wearing glasses when I used the headset, and almost everything looked clear but I could not read any of the posters on the wall*.” These critiques underscore the need for future iterations of the VR platform.

## IV. Discussion

Effective trauma training is pivotal for accurate and timely interventions, especially given the disparities observed globally.[28] However, the high costs of current courses limit trauma education dissemination.[12,13] Beyond the challenges of securing mannequins and instructors, current training recommendations for pediatric trauma are simply unfeasible.[29] For example, the American College of Surgeons’ *Resources for Optimal Care of the Injured Patient* of 2014 has recommended at least 16 hours of pediatric trauma training yearly for all healthcare providers caring for injured children. [29] However, even the Advanced Trauma Life Support (ATLS) course, the world’s most successful trauma course offered in 86 countries, has only been completed by 6% of the world’s physicians over the past 14 years.[30–32] Based on this scenario, we estimate that training four-member teams in every trauma center to treat the 60 million children who visit hospitals yearly[1,29,33–35] could require an impossible investment of 1.2 billion US dollars in pediatric trauma education annually.

Furthermore, the potentially immersive nature of VR, some with realistic scenarios, stands to enrich patient interactions and encourage collaboration among team members. VR may help to authentically replicate the stress factors of a hospital environment, including the presence of relatives, ambient noise, and interdepartmental dynamics. [36,37] Finally, VR platforms can provide automated feedback, such as a checklist of completed tasks and the time taken to perform them. Users’ recall following stressful events can be unreliable.[38] Having a generated list that details tasks performed—or not performed—by users, along with the timing of each, can assist instructors in guiding the debriefing process toward areas of weakness. Additionally, the availability of this automated record decreases the need for instructors to take notes during the session.[38]

Participants highlighted the benefit of this VR platform in enhancing non-technical skills over technical skills. While limited haptic feedback in VR has been noted as a barrier to practicing technical skills before, some authors disagree with this argument.[39–42] They underpin that successful technical tasks often hinge on decision-making processes, including selecting the correct equipment and identifying anatomical landmarks.[39–42] Given that decision-making is a critical component of non-technical skills and can be simulated in VR, the platform may offer substantial benefits even for technical skills.

In our opinion, to benefit most from the platform, it is crucial to define the learning objectives clearly, and identify target users *before* creating any virtual scenario. Our primary goal was to augment the capabilities of established trauma teams. For these experienced providers, technical skill acquisition (for example, inserting a chest tube) is less of an issue, allowing them to concentrate on other learning facets where this virtual platform has more benefits, such as encouraging communication and collaboration between users in a stressful environment.

In our study, 27% of the users reported significant sickness, which raised initial concerns among the research team. However, follow-up discussions with participants indicated that two (18%) were not wearing their usual prescription glasses, which can cause eye strain issues. Other participants believed they could discern all details, such as small text on a virtual flier. They reported blurry vision or difficulty focusing on the cybersickness survey. Yet, these issues are known limitations of the current headset’s display resolution and optics rather than project-virtual simulation issues. Furthermore, the interpupillary distance (IPD) setting within the headset itself was not calibrated for each user, which may cause users with an IPD further from the norm to encounter a blurry stereoscopic view, contributing to dizziness and eye fatigue.[43]

The feedback obtained will be instrumental in refining our tutorial to clarify the platform’s limitations and to instruct users to take the necessary time to adjust the headset for optimal comfort. Users will be advised to take breaks where required, and IPD will be calibrated for each user individually before their virtual simulation is conducted. Future test headsets will utilize newer display technology with improved resolution and optics to further alleviate issues. Further evaluations will be conducted to ensure the cybersickness level after these modifications.

### Limitations and Future Directions

A significant limitation of this preliminary evaluation is the reliance on data from a small cohort, which, despite its diverse backgrounds, works or studies at the same university health center. Nonetheless, we consider this initial phase critical as a precursor to more resource-intensive studies. Future phases will further assess acceptance, cybersickness, and efficacy across a broader demographic sample through different trials in different centers. This includes settings with lower internet connection quality where simulation-based training alternatives might be scarce.

The study was conducted in Canada, with a relatively young generation of physicians, and approximately one-third of the participants had prior exposure to or experience with virtual simulation. It is recognized that previous experience with virtual simulation may confer an advantage, potentially contributing to enhanced outcomes.[44] A sub-analysis of different cohorts—those with and without experience—may be essential to fully comprehend the impact of this technology on users who are not accustomed to it.

For the purposes of refining the virtual world, we also aim to conduct a more detailed analysis of its features, including the validity of the automatic feedback provided.

## V. Conclusion

Our preliminary assessment of a VR platform for pediatric trauma education has yielded promising results. Users expressed satisfaction with the realism of the virtual trauma bay, the virtual patient, and the equipment. The platform was deemed particularly effective for enhancing non-technical skills over technical skills, with potential benefits extending across a broad spectrum of users, from medical students to experienced staff. Finally, most participants concurred that the tested VR platform offers a more immersive simulation training experience than high-fidelity mannequins.

Nonetheless, moderate cybersickness was reported by almost one-third of the participants. Even though this may be attributed to hardware limitations, such as the inability to render fine details, this concern warrants careful investigation. Given that VR can help address crucial gaps in pediatric trauma education—namely, accessibility and non-technical skills training, these initial findings encourage subsequent trials to compare VR simulation and high-fidelity mannequins.

## Data Availability

All data produced in the present study are available upon reasonable request to the authors

